# Liver iron levels are associated with *HFE*-hemochromatosis genotype, diet, adiposity, and disease in the UK Biobank

**DOI:** 10.1101/2025.04.29.25322001

**Authors:** Mitchell R Lucas, Luke C Pilling, João Delgado, Daniel S. Williamson, Jeremy D Shearman, Katharine Hutchison, Janice L Atkins

**Affiliations:** Department of Clinical and Biomedical Sciences, Faculty of Health and Life Sciences, University of Exeter, Exeter, UK; Department of Health and Care Professions, Faculty of Health and Life Sciences, University of Exeter, Exeter, UK; Department of Gastroenterology, South Warwickshire NHS Foundation Trust, UK; Department of Gastroenterology, Royal Devon University Healthcare NHS Foundation Trust, Exeter, Devon, UK

## Abstract

**Introduction:** *HFE* genetic variants, especially C282Y homozygosity (C282Y+/+), can increase systemic iron and cause hemochromatosis, though expression varies. Excess iron can lead to liver disease/cancer, yet factors influencing liver iron beyond *HFE* genotype remain unclear. We investigated genetic/environmental factors influencing liver iron, including *HFE* genotype and hemochromatosis diagnosis.

**Methods:** We analysed 37,287 European ancestry UK Biobank participants (mean age 64.1, SD 7.6) with *HFE* genotypes and MRI-estimated liver iron concentrations (MRLIC). Linear regression assessed MRLIC associations with genetic and environmental factors, adjusting for age, sex, and genetic covariates.

**Results:** Mean MRLIC was highest in undiagnosed C282Y+/+ males and females (2.56 and 2.31mg/g) vs. diagnosed (1.23 & 1.51mg/g, p=0.0001 & 0.0004). Other *HFE* genotypes had nominal increases vs. those without *HFE* genetic variants. Higher MRLIC was associated with higher alcohol intake (β=0.11, 95%CI: 0.09–0.11, p=6.0×10^−128^; >30 vs. 1–14 units/week), frequent red/processed meat consumption (β=0.08, 95%CI: 0.07–0.09, p=3.7×10^−54^; ≥3 times/week vs. none), high waist-height ratio (β=0.01, 95%CI: 0.006–0.02, p=6.4×10^−5^) and genetically predicted transferrin saturation (β=0.22, 95%CI: 0.19– 0.26, p=3.8×10^−46^). Lower MRLIC was associated with underweight BMI (β=−0.06, 95%CI: −0.09 – −0.03, p=1.1×10^−4^), proton pump inhibitor use (β=−0.03, 95%CI: −0.04 – −0.03, p=3.5×10^−17^), and ≥4 cups/day of tea (β=−0.01, 95%CI: −0.02 – −0.003, p=0.005 vs. non-drinkers). Among C282Y+/+ individuals (n=91), tea ≥4 cups/day was associated with lower MRLIC (β=−0.99, 95%CI: −1.89 – −0.08, p=0.03).

**Conclusion:** Undiagnosed C282Y+/+ individuals had excess liver iron vs. diagnosed, likely due to treatment. Genetic and environmental factors influence liver iron beyond C282Y+/+. Tailored lifestyle advice could benefit those at risk of hemochromatosis.

## Introduction

Hemochromatosis is the most common genetic disorder among people of Northern European descent.^1^ The condition is characterized by iron accumulation, primarily in the liver.^2^ It can be associated with severe clinical outcomes, including liver disease, liver cancer, joint debilitation, neurodegenerative diseases and diabetes.^3–7^ The strongest risk factor is the *HFE* C282Y genetic variant, with 1 in 150 individuals of Northern European ancestry being C282Y homozygous (C282Y+/+).^8^ While C282Y+/+ exhibit the highest penetrance among *HFE* genotypes, such as elevated transferrin saturation (TSAT) and serum ferritin (SF),^9^ not all go on to be diagnosed with hemochromatosis or associated clinical manifestations.^3^ Other *HFE* C282Y and H63D genotype combinations (such as C282Y-H63D compound heterozygotes) have far lower penetrance.^9^ The clinical significance of *HFE* C282Y and H63D variants in iron loading and related outcomes remains unclear.

Genetics and environmental factors influence iron status within the body.^10,11^ Alcohol consumption exacerbates iron accumulation, particularly in C282Y+/+.^12^ Central adiposity has been correlated with lower levels of liver iron^13^, though findings are mixed.^14^ However, interactions between genetic and other modifiers such as vitamin/mineral supplement intake, dietary habits, lifestyle, and other factors have yet to be studied in large community-based. Understanding how genetics and lifestyle influence hemochromatosis-related phenotypes may help to explain the varied clinical penetrance observed in *HFE* genotypes.

To address these research gaps, we analyzed data from the UK Biobank, the largest available cohort of *HFE* genotyped participants with magnetic resonance imaging (MRI) estimated liver iron concentrations (MRLIC), to investigate the genetic and environmental factors influencing liver iron status. MRI is a well-established and highly sensitive method for detecting iron in organs, particularly the liver, making it an effective tool for measuring liver iron levels.^15^ Given the low levels of iron accumulated within reticuloendothelial cells in hemochromatosis, it has been inferred that a liver-to-spleen iron ratio (LSIR) may increase the diagnostic specificity of MRI.^16,17^ Using MRI-derived tissue iron data (liver and spleen) from the UK Biobank, we aim to identify the strongest predictors of iron deposition. First, we compared MRLIC and LSIR by *HFE* genotype, undiagnosed with hemochromatosis, to assess penetrance across each genotype group. Second, we examined genetic and environmental factors associated with iron loading. Finally, we compared mean MRLIC and LSIR across *HFE* genotypes diagnosed with hemochromatosis.

## Methods

### Study Population

The UK Biobank includes over 500,000 community volunteers aged 37 to 73 years at baseline (2006-2010) from 22 assessment centres across England, Scotland and Wales. Among this cohort, 451,270 participants were genetically similar to the 1000 Genomes project European reference population,^18^ with *HFE* C282Y (rs1800562) and *HFE* H63D (rs1799945) genotypes from whole exome sequencing (methods by Regeneron).^19^ The UK Biobank has approval from the North-West Multi-Centre Research Ethics Committee, and participants gave written informed consent before the study (Research Ethics Committee reference 11/NW/0382). The protocol conformed to the Declaration of Helsinki. Access to the UK Biobank was granted under application number 14631. Participants were informed of baseline health-related findings, but consent did not cover notification of incidental findings from subsequent analyses, including genotyping result. To protect participant privacy, the UK Biobank require that data or aggregate statistics corresponding to fewer than 5 participants cannot be published, nor can data or statistics be reported that allow a participant count of less than 5 to be derived.^20^

### Baseline assessment (2006-2010)

Socioeconomic status was recorded at baseline using the Townsend deprivation index, which integrates unemployment, lack of car ownership, absence of homeownership, and household overcrowding within a participant’s region of residence.^21^ A greater Townsend deprivation index indicates an increased material disadvantage. Measures of blood glucose were measured using the hemoglobin A1c (HbA1c) test, measured by high-performance liquid chromatography analysis on a Bio-Rad VARIANT II Turbo.^22^ Total cholesterol and triglycerides were measured using a Beckman Coulter AU5800.^22^

A previously conducted genome-wide association study (GWAS) was performed on four key iron-related biomarkers, utilizing data from the Trøndelag Health Study (HUNT), the Michigan Genomics Initiative (MGI), and the SardiNIA study.^23^ This research identified 123 genetic loci linked to iron homeostasis and their impact on all-cause mortality.^23^ Genetic variants associated with TSAT and SF from these cohorts (excluding the *HFE* gene) were used to create polygenic risk scores (PGS), which determined participants genetic likelihood of having high TSAT and SF separately.

### Imaging visit (2014-2020)

A subset of 37,287 UK Biobank European ancestry participants of (17,780 males; 19,507 females) underwent abdominal MRI between 2014 and 2020; data was available on liver iron (field: 40060 n=37,287), spleen iron (field: 21170, n=29,411), pancreas iron (field: 21091, n=25,907), and liver fat proton density fat fraction (PDFF) (field: 40061, n=36,420).^24^ MRI liver iron data was also available on 2,859 participants of non-European ancestry: spleen iron n=2,332; pancreas iron n=2,016; liver PDFF n=2,060. Clinical cut-offs previously determined for liver iron levels were <1.8 mg/g (normal), ≥1.8 to <3.2 mg/g (borderline excess), ≥3.2 to <7.0 mg/g (mild iron overload), ≥7.0 to 15.0 mg/g (moderate iron overload) and >15.0 mg/g (severe iron overload).^27^ LSIR was calculated by dividing liver iron (mg/g) by spleen iron (mg/g). A high LSIR often suggests hemochromatosis-related iron overload. In contrast, a low LSIR is typically associated with secondary iron overload conditions, such as those resulting from blood transfusions or iron-loading anemias.^16,17^

At the time of the MRI visit, participants had their weight, height, waist circumference (WC), and hip circumference (HC) measured. We calculated body mass index (BMI) as weight (kg) divided by height (m^2^), waist-hip-ratio (WHR) by dividing WC (cm) by HC (cm), and waist-height ratio (WHtR) by diving WC (cm) by height (cm). Questionnaires collected information on doctor-diagnosed conditions (including hemochromatosis, hypertension, type 1 and 2 diabetes and viral hepatitis), vitamin and mineral supplement intake, medications, dietary intake, smoking status, alcohol intake, physical activity and education. Alcohol intake was categorized by the number of units drank per week groups into ‘0 units per week’ ‘1-14 units per week’ (reference group, based on UK national recommendations)^25^ ‘15-29 units per week’ and ‘over 30 units per week’. Using data collected on daily tea drinking, which was derived from being asked “How many cups of tea do you drink each day? (Including black and green tea)”, we categorized data into ‘0 cups’, ‘1-3 cups’ and ‘4 or more cups’. We calculated the number of participants taking iron supplements and vitamin C supplements using data collected from participants being asked “Do you regularly take any of the following?”. Vitamin C is of interest due to its relationship with increased iron absorption.^26^ Additionally, we calculated how many participants took proton pump inhibitors (PPI) from participants being asked “In the touch screen you said you are taking regular prescription medications. Can you now tell me what these are?”. Smoking status was defined as ‘current smoker’ based on subjects being asked “Do you smoke tobacco now? and grouped in a binary variable of 0=‘No’ and 1=‘Yes, on most or all days’ and ‘Only occasionally’. Participants were asked how often they ate processed meat, lamb/mutton, pork, and beef. For each type of meat, participants could choose from the following options: ‘Never’, ‘Less than once a week’, ‘Once a week’, ‘2-4 times a week’, ‘5-6 times a week’, or ‘Once or more daily’. We provided a value for each respective response on meat consumption: (Never=0) (Less than once a week=0.5) (Once a week=1) (2-4 times a week=3) (>4 times a week=5.5) then derived a summed total weekly consumption of red /processed meat and recorded as follow ‘0 times/week’ (reference group) and ‘0.1-2.9 times/week’, and ‘≥3.0 times/week’.^27^ Highest educational qualification was ranked as: 0=none, 1=CSEs (Certificate of Secondary Education), 2=GCSEs/O-levels (General Certificate of Secondary Education to age 16), 3=A-levels/NVQ/HND/HNC (further education after age 16), 4=professional qualification, and 5=college/university degree.

### Ascertainment of diagnoses

Ascertainment of disease diagnoses was derived from baseline questionnaires plus International Classification of Diseases, 10th Revision (ICD-10) coded hospital inpatient data (National Health Service Hospital Episode Statistics) from 1996 to the time of the MRI scans (2014-2020). Diseases included hemochromatosis (ICD-10 code E83.1), viral hepatitis (B15, B16, B17, B18, B19, K73), hypertension (I10, I11, I12, I13, I14, I15), diabetes (type 1 and 2 [E10; E11]), liver fibrosis and cirrhosis (K74), and any liver disease (K70, K71, K72, K73, K74, K75, K76, K77)

### Statistical Analysis

Descriptive statistics were used to summarise the data in 37,287 participants with (n=58) and without hemochromatosis (n=37,229). We used the Kruskal-Wallis test to assess differences in mean MRI liver iron concentration (MRLIC), LSIR, liver PDFF, and pancreas iron between *HFE* genotype groups. Normality was checked via Q-Q plots and Shapiro-Wilk tests; data remained non-normal despite log transformations. Logistic regression models were used to compare differences in proportions with borderline excess liver iron and mild iron overload between *HFE* genotype groups.

Participants diagnosed with hemochromatosis were excluded from further regression analyses to avoid bias from potentially receiving ongoing iron monitoring and treatment. Linear regression models examined associations between MRLIC and genetic factors (PGS for TSAT and SF) and environmental factors (WHR, BMI, WHtR, alcohol intake, smoking, diet, supplements, PPI use, blood biomarkers, and disease diagnoses). Analyses were conducted within European ancestry participants regardless of genotype (n=37,229), participants without C282Y/H63D variants (n=22,388), C282Y+/+ specifically (n=91), and a combined group of other *HFE* genotypes without hemochromatosis (n=14,750, including H63D+/-, H63D+/+, C282Y+/H63D+, and C282Y+/-). Additionally, analyses were performed among non-European ancestry participants (n=2,859). Models were adjusted for age, sex, and 10 genetic principal components (PCs); adjusting for PCs reduces bias due to population stratification.^28^ Models were additionally adjusted for covariates known to be associated with iron status:^16,29^ assessment center, red/processed meat consumption, WHR, alcohol intake, PPI use, education, socioeconomic status, smoking, hepatitis, type 2 diabetes, tea drinking, and iron and vitamin C supplements.

Cox proportional hazards models estimated the associations between MRLIC and risk of liver-related incident outcomes (liver disease, liver fibrosis or cirrhosis, and hemochromatosis), excluding prevalent disease. Models were adjusted for age, sex, and 10 PCs, within participants of European ancestry (n=37,229), without C282Y/H63D variants (n=22,388), and C282Y+/+ (n=91). Sensitivity analyses excluded prevalent anemia cases and repeated initial models by adjusting for potential confounders including alcohol intake, smoking, BMI, and prevalent comorbidities: type 2 diabetes, any viral hepatitis, alcoholic liver disease, and non-alcoholic fatty liver disease (NAFLD). We also assessed competing risks with outcomes including mortality, liver cancer, and hepatitis. Cox proportional hazards assumptions were confirmed with ‘estat phtest’. Bonferroni correction for 27 regression models set significance at p<0.002. Analyses were conducted using Stata v18.0; figures were created using R.

## Results

### Males

We analyzed 17,780 males (mean age 64.8 years, SD 7.7), including 45 C282Y+/+ individuals, of which 22 were diagnosed with hemochromatosis (48.89%). Undiagnosed C282Y+/+ males had the highest mean MRI liver iron concentration (MRLIC) at 2.56 mg/g (SD 1.4), compared to 1.23 mg/g (SD 0.2) in those with a hemochromatosis diagnosis (p=0.0001). Similarly, MRLIC differed by hemochromatosis status in male C282Y+/H63D+, with values of 1.63 mg/g (SD 0.6) vs. 1.27 mg/g (SD 0.1) (p=0.02). Other *HFE* genotypes had nominal increases in MRLIC compared to those without *HFE* genetic variants (Table 1; Figure 1).

**Table 1.** Characteristics of male UK Biobank participants of European Ancestry, by C282Y and H63D genotypes.

|  | Males |  |  |  |  |  | Total | p-value |
| --- | --- | --- | --- | --- | --- | --- | --- | --- |
|  | No C282Y<br>or H63D<br>variants | H63D+/- | H63D+/+ | C282Y+/<br>H63D+ | C282Y+/- | C282Y+/+ |  |  |
| **No hemochromatosis diagnosis, n (%)** | 10,702<br>(99.95) | 4,125 (99.95) | 362 (97.72) | 401 (97.33) | 2,125 (99.95) | 23 (51.11) | 17,738 | . |
| Liver iron (mg/g), mean (SD) | 1.27 (0.2) | 1.31 (0.3) | 1.44 (0.5) | 1.63 (0.6) | 1.34 (0.3) | 2.56 (1.4) | 1.30 (0.3) | 0.0001 |
| Liver iron groups: |  |  |  |  |  |  |  | 0.0001 |
| Borderline excess liver iron ($\geq 1.8$ to $< 3.2$ mg/g), n (%) | 325 (3.04) | 213 (5.16) | 47 (12.98) | 109 (27.18) | 149 (7.01) | <5 | 847 (4.78) | . |
| Mild iron overload ($\geq 3.2$ to $< 7$ mg/g), n (%) | <5 | 7 (0.17) | 6 (1.66) | 12 (2.99) | <5 | 10 (43.48) | 41 (0.23) | . |
| Liver-to-spleen ratio, mean (SD) | 4.16 (1.0) | 4.33 (1.1) | 4.72 (1.7) | 5.50 (2.2) | 4.36 (1.2) | 9.26 (4.9) | 4.27 (1.1) | 0.0001 |
| Liver PDFF (%), mean (SD) | 5.52 (5.1) | 5.68 (5.2) | 5.82 (5.5) | 5.88 (5.6) | 5.76 (5.4) | 5.68 (3.2) | 5.6 (5.2) | 0.02 |
| Pancreas iron (mg/g), mean (SD) | 0.75 (0.1) | 0.75 (0.1) | 0.75 (0.1) | 0.74 (0.1) | 0.74 (0.1) | 0.74 (0.1) | 0.75 (0.1) | 0.71 |
| **Hemochromatosis diagnosis, n (%)** | 5 (0.05) | <5 | <5 | 11 (2.67) | <5 | 22 (48.89) | 42 | . |
| Liver iron (mg/g), mean (SD) | 1.66 (1.2) | - | - | 1.27 (0.1) | - | 1.23 (0.2) | 1.30 (0.4) | 0.48 |
| Liver iron groups: |  |  |  |  |  |  |  | 0.18 |
| Borderline excess liver iron ($\geq 1.8$ to $< 3.2$ mg/g), n (%) | 0 (0.00) | - | - | 0 (0.00) | - | 0 (0.00) | 0 (0.00) | . |
| Mild iron overload ($\geq 3.2$ to $< 7$ mg/g), n (%) | <5 | - | - | 0 (0.00) | - | 0 (0.00) | <5 | . |
| Liver-to-spleen ratio, mean (SD) | 7.24 (5.6) | - | - | 4.39 (0.6) | - | 4.18 (1.0) | 4.51 (1.9) | 0.69 |
| Liver PDFF (%), mean (SD) | 10.18 (13.5) | - | - | 10.53 (9.3) | - | 6.32 (6.4) | 7.89 (8.0) | 0.40 |
| Pancreas iron (mg/g), mean (SD) | 0.65 (0.1) | - | - | 0.78 (0.1) | - | 0.75 (0.1) | 0.75 (0.1) | 0.14 |

Male participants (n=17,780) genetically similar to the 1000 Genomes Project European Ancestry superpopulation ('EUR-like')<sup>18</sup> with available *HFE* genotypic data and MRI liver iron imaging data in the UK Biobank. Numbers are presented as mean (SD) for continuous variables and n (%) for categorical variables. Abbreviations: mg/g, milligrams per gram; MRI, magnetic resonance imaging; PDFF, proton density fat fraction; SD, standard deviation. "--="not reported due to too few participants.

**Figure 1.**
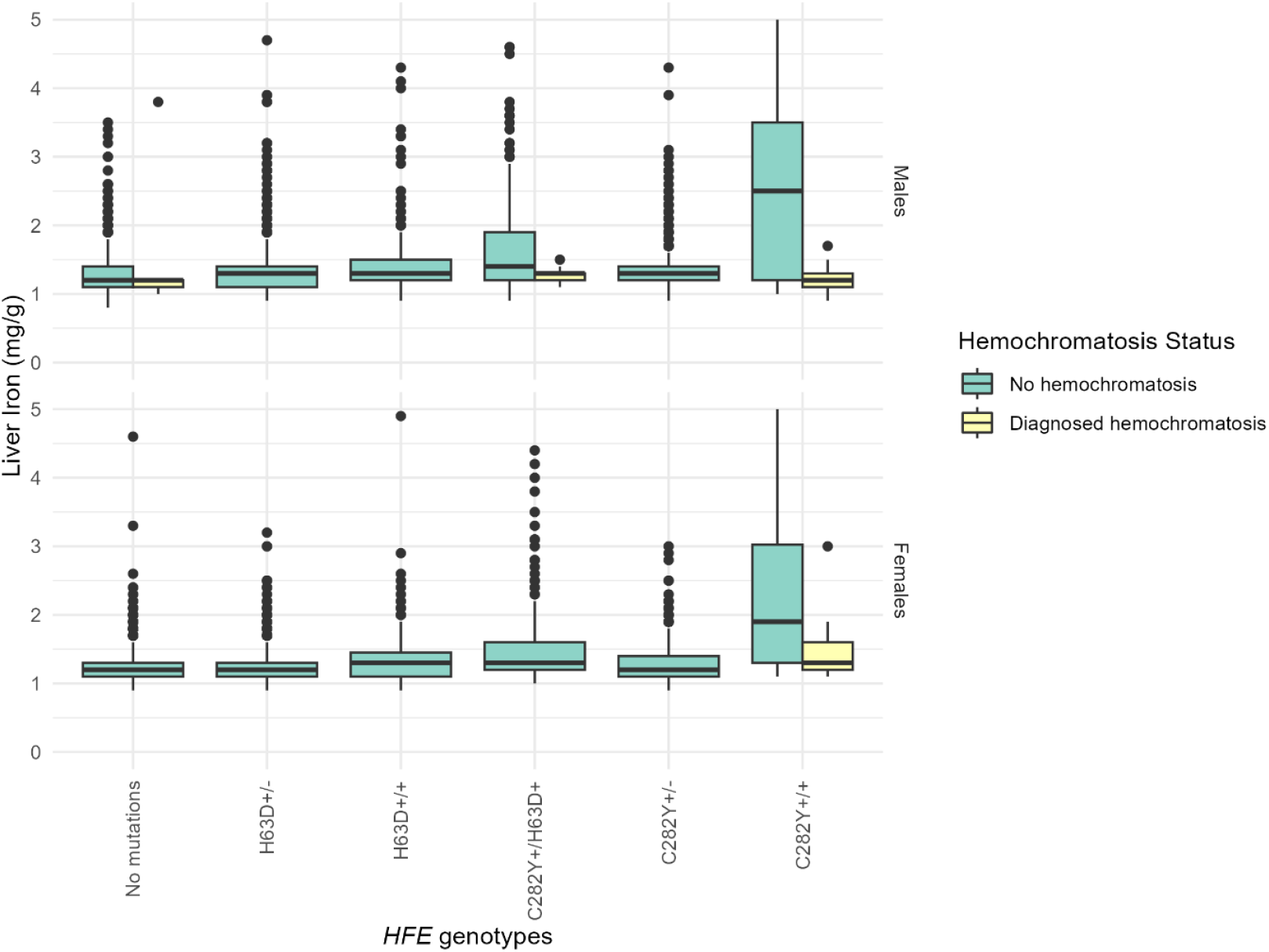
Distribution of liver iron by *HFE* genotype and hemochromatosis diagnosis. Distribution of MRI liver iron (male n=17,780; female n=19,507) by *HFE* genotypes, stratified by hemochromatosis diagnosis, in the UK Biobank. Abbreviations: mg/g, milligrams per gram; MRI, magnetic resonance imaging.

Undiagnosed C282Y+/+ also had the highest prevalence of mild iron overload (≥3.2 to <7 mg/g) (n=10, 43.48%). Borderline levels of excess liver iron (≥1.8 to <3.2 mg/g) were most common in undiagnosed male C282Y+/H63D+ (n=109, 27.18%), compared to 3.04% in males without C282Y and H63D variants. Undiagnosed C282Y+/+ males had the highest mean LSIR at 9.26 (SD 4.9), followed by C282Y+/H63D+ males at 5.50 (SD 2.2), compared to those without *HFE* variants (4.16, SD 1.0). Other *HFE* genotypes showed a comparable LSIR distribution to males without *HFE* variants undiagnosed with hemochromatosis. There was no significant difference in liver PDFF % between C282Y+/+ individuals without and with hemochromatosis, with values of 5.68% and 6.32%, respectively (p=0.69); similar results were observed in C282Y+/H63D+ individuals (5.88% vs. 10.53%, p=0.15). Pancreas iron levels did not differ significantly in males between *HFE* genotypes (p=0.71 for those without a diagnosis; p=0.14 for those with a diagnosis) (Table 1; eFigures 1-3).

### Females

We analyzed 19,507 females (mean age 63.5 years, SD 7.5), including 80 C282Y+/+ individuals, of which 12 were diagnosed with hemochromatosis (15.00%). Mean liver iron concentration was highest in female C282Y+/+ individuals without a hemochromatosis diagnosis at 2.31 mg/g (SD 1.2), compared to 1.51 mg/g (SD 0.5) in those with a hemochromatosis diagnosis (p=0.0004). Other *HFE* genotypes had nominal increases in MRLIC compared to those without *HFE* genetic variants (Table 2; Figure 1).

**Table 2.** Characteristics of female UK Biobank participants of European Ancestry, by C282Y and H63D genotypes.

|  | Females |  |  |  |  |  | Total | p-value |
| --- | --- | --- | --- | --- | --- | --- | --- | --- |
|  | No C282Y<br>or H63D<br>variants | H63D+/- | H63D+/+ | C282Y+/<br>H63D+ | C282Y+/- | C282Y +/+ |  |  |
| **No hemochromatosis diagnosis, n (%)** | 11,686 (100) | 4,624<br>(99.96) | 443<br>(99.77) | 408 (100) | 2,262<br>(99.96) | 68 (85.00) | 19,491 | . |
| Liver iron (mg/g), mean (SD) | 1.22 (0.2) | 1.25 (0.2) | 1.33 (0.3) | 1.47 (0.5) | 1.27 (0.2) | 2.31 (1.2) | 1.25 (0.2) | 0.0001 |
| Liver iron groups: |  |  |  |  |  |  |  | 0.0001 |
| Borderline excess liver iron ($\geq 1.8$ to $< 3.2$ mg/g), n (%) | 188 (1.61) | 118 (2.55) | 30 (6.77) | 63 (15.44) | 85 (3.76) | 23 (33.82) | 507 (2.60) | . |
| Mild iron overload ($\geq 3.2$ to $< 7$ mg/g), n (%) | <5 | <5 | <5 | 6 (1.47) | 0 (0.00) | 16 (23.53) | 26 (0.13) | . |
| Liver-to-spleen ratio, mean (SD) | 4.19 (0.8) | 4.28 (0.9) | 4.53 (1.3) | 5.01 (1.9) | 4.34 (0.9) | 8.17 (4.2) | 4.27 (1.0) | 0.0001 |
| Liver PDFF (%), mean (SD) | 4.22 (4.6) | 4.21 (4.5) | 4.45 (4.5) | 4.36 (4.6) | 4.34 (4.7) | 4.61 (4.3) | 4.24 (4.6) | 0.02 |
| Pancreas iron (mg/g), mean (SD) | 0.80 (0.1) | 0.80 (0.1) | 0.81 (0.1) | 0.80 (0.1) | 0.80 (0.1) | 0.80 (0.1) | 0.80 (0.1) | 0.83 |
| **Hemochromatosis diagnosis, n (%)** | 0 (0.00) | <5 | <5 | 0 (0.00) | <5 | 12 (15.00) | 16 | . |
| Liver iron (mg/g), mean (SD) | 0 (0.0) | - | - | 0 (0.0) | - | 1.51 (0.5) | 1.46 (0.5) | 0.49 |
| Liver iron groups: |  |  |  |  |  |  |  | 0.86 |
| Borderline excess liver iron ($\geq 1.8$ to $< 3.2$ mg/g), n (%) | 0 (0.00) | - | - | 0 (0.00) | - | 0 (0.00) | <5 | . |
| Mild iron overload ($\geq 3.2$ to $< 7$ mg/g), n (%) | 0 (0.00) | - | - | 0 (0.00) | - | <5 | <5 | . |
| Liver-to-spleen ratio, mean (SD) | 0 (0.0) | - | - | 0 (0.0) | - | 4.82 (2.5) | 4.75 (2.2) | 0.34 |
| Liver PDFF (%), mean (SD) | 0 (0.0) | - | - | 0 (0.0) | - | 7.34 (9.3) | 7.6 (8.9) | 0.31 |
| Pancreas iron (mg/g), mean (SD) | 0 (0.0) | - | - | 0 (0.0) | - | 0.88 (0.1) | 0.85 (0.1) | 0.33 |

Female participants (n=19,507) genetically similar to the 1000 Genomes Project European Ancestry superpopulation ('EUR-like')<sup>18</sup> with available *HFE* genotypic data and MRI liver iron imaging data in the UK Biobank. Numbers are presented as mean (SD) for continuous variables and n (%) for categorical variables. Abbreviations: mg/g, milligrams per gram; MRI, magnetic resonance imaging; PDFF, proton density fat fraction; SD, standard deviation. "--" = not reported due to too few participants.

The prevalence of mild iron overload (≥3.2 to <7 mg/g) was also highest in undiagnosed female C282Y+/+ (n=16, 23.53%). Borderline levels of excess liver iron (≥1.8 to <3.2 mg/g) were most common in undiagnosed female C282Y+/+ (n=23, 33.82%) and C282Y+/H63D+ (n=63, 15.44%). Undiagnosed C282Y+/+ females had the highest LSIR at 8.17 (SD 4.2), followed by C282Y+/H63D+ females at 5.01 (SD 1.9), while those without *HFE* variants had the lowest LSIR at 4.19 (SD 0.8). LSIR in other undiagnosed female *HFE* genotypes did not differ compared to those without *HFE* variants. There was no significant difference in liver PDFF % between C282Y+/+ individuals without and with hemochromatosis, with values of 4.61% and 7.34%, respectively (p=0.34). Pancreas iron did not differ significantly in female between *HFE* genotype groups, for those undiagnosed (p=0.83) and diagnosed (p=0.33) (Table 2; eFigures 1-3).

### Associations between genetic and environmental factors with magnetic resonance liver iron concentration in participants undiagnosed with hemochromatosis

Within 37,229 male and female participants of European ancestry undiagnosed with hemochromatosis, PGS for TSAT and SF were positively associated with MRLIC (β=0.22, 95% CI: 0.19 to 0.26, p=3.8×10^−46^ and β=0.18, 95% CI: 0.15 to 0.21, p=2.1×10^−27^, respectively). Alcohol intake showed a positive dose-response relationship with MRLIC, with consumption of more than 30 units per week showing the strongest association (β=0.11, 95% CI: 0.09 to 0.11, p=6.0×10^−128^) compared to those drinking 1–14 units per week. Red/processed meat consumption exhibited a positive dose-response relationship, with intake of 3 or more times per week most strongly associated with MRLIC (β=0.08, 95% CI: 0.07 to 0.09, p=3.7×10^−54^), compared to those consuming no portions per week. Being a current smoker was also positively associated with MRLIC (β=0.03, 95% CI: 0.02 to 0.04, p=5.1×10^−5^), as was a high WHtR (β=0.01, 95% CI: 0.006 to 0.02, p=6.4×10^−5^).

In contrast, being diagnosed with type 1 or 2 diabetes was negatively associated with MRLIC (β=−0.08, 95% CI: −0.12 to −0.03, p=9.9×10^−4^ and β=−0.07, 95% CI: −0.08 to −0.05, p=8.2×10^−17^, respectively). Similarly, having an underweight BMI was associated with lower MRLIC (β=−0.06, 95% CI: −0.09 to −0.03, p=0.0001) compared to a normal BMI, whereas an overweight BMI was associated with higher MRLIC (β=0.02, 95% CI: 0.02 to 0.03, p=4.3×10^−15^).

PPI medication use was also negatively associated with liver iron levels (β=−0.03, 95% CI: −0.04 to −0.03, p=3.5×10^−17^). Tea drinking was negatively associated with MRLIC among those who consumed 4 or more cups daily (β=−0.01, 95% CI: −0.02 to −0.003, p=0.005) compared to non-tea drinkers; however, this association became non-significant after correction for multiple testing (eTables 1 and 2; Figure 2).

**Figure 2:**
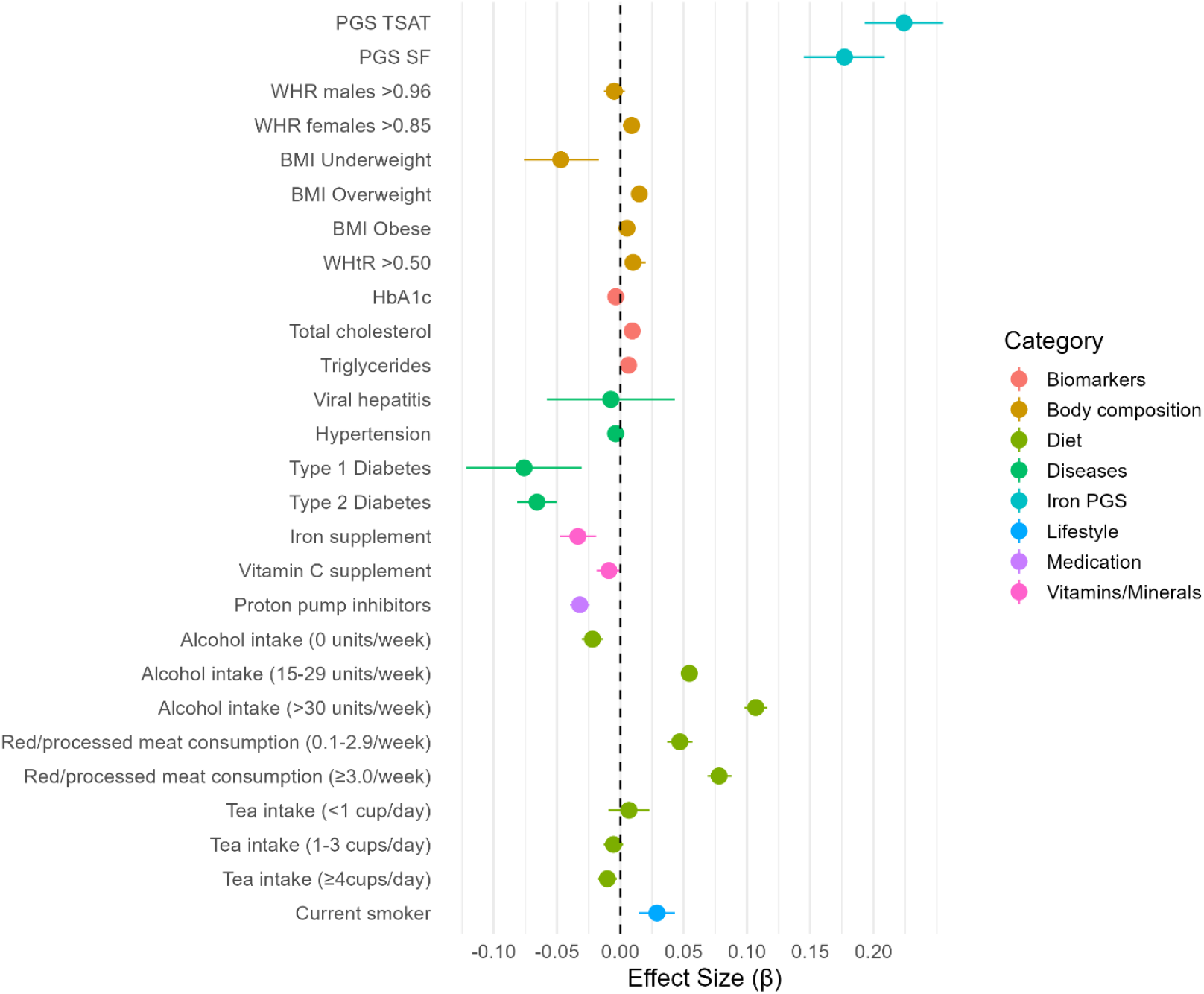
Associations between environmental and genetic variables and magnetic resonance liver iron concentration in UK Biobank participants of European Ancestry undiagnosed with hemochromatosis. Forest plot of beta coefficients and 95% confidence intervals for MRLIC data, adjusted for age, sex, and 10 genetic principal components. The plot shows associations of iron polygenic score (PGS: TSAT and SF), body composition, blood biomarkers, diseases, vitamins/minerals, medication, diet, and lifestyle with magnetic resonance liver iron concentration in EUR-like^18^ participants without a hemochromatosis diagnosis (n=37,229). Reference groups include normal BMI for BMI body composition, 1–14 units/week for alcohol intake, 0 times/week for meat consumption, and 0 cups/day for tea intake. Abbreviations: BMI, body mass index; MRLIC, MRI liver iron concentration; PGS, polygenic score; SF, serum ferritin; TSAT, transferrin saturation; WHR, waist-to-hip ratio; WHtR, waist-to-height ratio.

After controlling for multiple testing, individuals with *HFE* variants (excluding C282Y homozygotes) (n=14,750) showed stronger associations with MRLIC for TSAT PGS, HbA1c, triglycerides, type 2 diabetes, proton pump inhibitor use, high alcohol and red/processed meat intake, and current smoking status compared to those without C282Y and H63D variants (n=22,388) (eTable 2). Among the 91 undiagnosed C282Y+/+ participants, MRLIC only showed a statistical negative association with tea drinking in those who consumed 1–3 cups daily (β=−0.91, 95% CI: −1.76 to −0.05, p=0.04) and 4 or more cups daily (β=−0.99, 95% CI: −1.90 to −0.09, p=0.03) compared to non-tea drinkers (eTable 2). However, this association did not remain significant after Bonferroni correction (p<0.002).

In non-EUR participants (n=2,859), SF PGS was associated with higher MRLIC (β=0.13, 95% CI: 0.02 to 0.25, p=0.03), while the TSAT PGS was not. Comparatively, in EUR participants, both TSAT and SF PGSs were significantly associated with MRLIC, with larger effect sizes. Alcohol and red/processed meat intake also showed significant positive, dose-response associations with MRLIC in non-EUR, but with slightly stronger effects in EURs. WHtR >0.50 was associated with higher MRLIC in both groups, though other body composition variables (underweight and overweight BMI) were only associated in EURs. Type 2 diabetes was inversely associated with MRLIC in both groups. Type 1 diabetes showed a positive association in non-EURs and a negative one in EURs. PPIs were not significantly associated with MRLIC in non-EURs (eTable 2).

In fully adjusted models, several associations observed in age, sex, and PC-adjusted analyses were no longer statistically significant (eTable 3). In contrast, several associations became statistically significant after full adjustment: red/processed meat intake ≥3 times/week was positively associated with liver iron in C282Y+/+ (β=1.13, 95% CI: 0.06–2.21, p=0.04), and frequent tea drinking (≥4 cups/day) was associated with lower liver iron in individuals without *HFE* variants (β=–0.009, 95% CI: –0.02 to –0.0008, p=0.03). However, none of the associations that became statistically significant in the fully adjusted models remained significant after correcting for multiple tests using the Bonferroni threshold.

### Associations between magnetic resonance liver iron concentration and risk of incident disease

Among individuals of European descent, undiagnosed with hemochromatosis at time of imaging, MRLIC was significantly associated with an increased risk of hemochromatosis (HR=5.40, 95% CI: 3.50 to 8.33, p=2.6×10^−14^). The association between MRLIC and incident diagnosis of liver fibrosis or cirrhosis was not significant after adjusting for multiple statistical testing (HR=0.10, 95% CI: 0.01 to 0.73, p=0.02) eTable 4). In competing risk models, with mortality, liver cancer, or hepatitis prior to the outcome, the association with liver fibrosis/cirrhosis remained non-significant(.

## Discussion

Hemochromatosis penetrance and clinical severity among *HFE* genotypes vary significantly. C282Y+/+ individuals typically express hemochromatosis-related phenotypes, such as raised TSAT and SF, and associated clinical outcomes more commonly than other *HFE* C282Y and H63D genotypes. However, there is no consensus on whether other *HFE* C282Y and H63D genotypes should be considered risk factors for hemochromatosis-related outcomes caused by iron loading. Our study highlights variations in liver iron by *HFE* genotype and hemochromatosis diagnosis (with the highest levels seen in undiagnosed C282Y+/+) as well as showing associations between genetic and environmental factors and liver iron levels. Our analysis is based on the largest community-based cohort with available data on *HFE* genotype and magnetic resonance liver iron measurements (n=37,287).

C282Y+/+ undiagnosed with hemochromatosis had the highest liver iron concentrations, with C282Y+/+ diagnosed with hemochromatosis having substantially lower liver iron; likely due to the effectiveness of hemochromatosis diagnosis and treatment in reducing liver iron. Other *HFE* genotypes had nominal increases in MRLIC compared to those without *HFE* genetic variants.

We found a strong, significant association between higher MRLIC and an increased risk of hemochromatosis, suggesting that MRLIC is a valuable tool for identifying iron overload. Despite excess liver iron being an established risk factor for liver fibrosis, cirrhosis, and liver cancer^30–32^, we found a nominal negative association between MRLIC and risk of liver fibrosis/cirrhosis. However, this association should be treated with caution due to the small sample size (n=39) and did not remain significant after Bonferroni correction.^30–32^

Alcohol can be a significant co-factor in hemochromatosis, with studies showing that alcohol consumption lowers hepcidin,^33,34^ and consuming 60g per day markedly increases the risk of liver fibrosis and cirrhosis.^12^ We demonstrate that alcohol consumption exceeding the recommended weekly limit (1-14 units)^25^ is associated with increased MRLIC in individuals of European ancestry, regardless of genotype, without a diagnosis of hemochromatosis. Fracanzani et al^35^ studied 452 Italian hemochromatosis patients and found smoking increased the risk of death (HR=2.1, 95% CI: 1.1 to 3.8; p=0.02) and hepatocellular carcinoma (HR=2.3, 95% CI: 1.2 to 2.7; p=0.01) compared to non-smokers. We observed an association between being a current smoker (vs non-smoker) and higher liver iron levels in European Ancestry participants of all genotypes without hemochromatosis; however, in our smaller sample of C282Y homozygotes (n=91) this association was not significant.

We previously identified that higher central adiposity in male (n=1,297) and female (n=1,602) C282Y+/+ individuals was associated with increased risks of liver complications.^36^ Pušeljić et al^13^ reported that fat distribution influences liver iron status in hemochromatosis patients (n=52), with intramuscular fat correlating positively (p=0.005, r_s_=0.382) and visceral subcutaneous fat correlating negatively (p<0.001, r_s_=−0.488) with liver iron overload. Powell et al^37^ reported BMI was independently associated with liver fibrosis in both light alcohol drinkers (OR=2.2, 95% CI: 1.2 to 4.2; p=0.01) and moderate-heavy alcohol drinkers (OR=3.6, 95% CI: 1.2 to 10.6; p=0.02). Our results support the notion that body composition is associated with liver iron status; we observed significant associations between overweight BMI and higher WHtR, with higher liver iron levels.

Iron overload increases the risk of diabetes,^38^ but here we observe a significant negative association between MRLIC and T1D/T2D. The effect a diagnosis of diabetes has on iron is unclear and although lifestyle changes impacting iron status are possible, population-based studies suggest that a diagnosis of type 2 diabetes does not result in drastic health behaviour changes.^39,40 41–44^The liver iron-diabetes association observed here therefore needs further investigation in prospective studies.

European^16^ and American^45^ liver disease guidelines recommend a healthy diet for hemochromatosis patients, avoiding iron and vitamin C supplements and iron-fortified foods, but provide limited advice on red meat. Salious et al^46^ found that in French C282Y+/+ hemochromatosis patients treated by phlebotomy (n=222), there was a weak association between consuming an iron-rich diet and greater iron overload. Rossi et al^47^ reported that higher consumption of red meat was associated with higher SF in males and females. Similarly, we observe a significant positive association between red/processed meat intake and MRLIC. Previously, a small clinical trial of 18 C282Y+/+ showed that black tea drinking with meals reduced intestinal iron absorption by around 70%,^48^ while another study showed no significant reduction in iron absorption from a single meal compared with water.^49^ Our results found a negative association between tea consumption and MRLIC in C282Y+/+ individuals without hemochromatosis in models adjusting for age, sex and PCs; this association did not remain significant after Bonferroni correction.

Over 25% of PPI users adhere to the medication for over a year;^50^ extended use can inhibit both non-heme and heme absorption.^51,52^ Among C282Y+/+ hemochromatosis patients assigned PPIs in a double-blind randomized controlled trial, the need for phlebotomies was significantly decreased vs. non-PPI users.^51^ Our findings support this literature, finding that PPI use was associated with lower liver iron levels within European ancestry participants regardless of genotype. Long-term PPI use can increase the risk of bone fractures,^53^ kidney disease,^54,55^ dementia,^56^ cardiovascular issues,^57^ and all-cause mortality.^57^ Therefore, despite potential benefits, current evidence does not support routine PPI use for hemochromatosis, and further research is needed to assess safety and efficacy.

Our findings highlight LSIR’s potential in diagnosing hemochromatosis in C282Y+/+ individuals, though we lacked data to track secondary iron overload patterns. C282Y+/+ males undiagnosed with hemochromatosis had the highest LSIR compared to other genotypes, consistent with iron loading within hepatocytes in hemochromatosis. In contrast, other genotypes showed balanced LSIR without signs of iron overload. While spleen iron measurements are less commonly used clinically than liver iron, a study by Sorokin et al^58^ showed their feasibility and relevance in iron metabolism, linking spleen iron to genetic variations in erythrocyte and macrophage-related genes.

### Strengths and limitations

We studied the largest available community-based cohort with *HFE* genotypesand available MRI data. Measures of serum ferritin and transferrin saturation were unavailable in the UK Biobank, but liver MRI is an established, non-invasive method for quantifying tissue iron, comparable to liver biopsy. UK Biobank participants were healthier than the general population at baseline.^59^ However, *HFE* allele frequencies were similar to other studies in the UK.^4^ Due to the absence of venesection treatment data, we used hemochromatosis diagnosis as a proxy, assuming diagnosed individuals are more likely to have received treatment, and excluded diagnosed individuals from the main analysis. Our study was cross-sectional in nature so the causality of associations cannot be inferred. The negative association of liver iron and tea consumption should be interpreted cautiously as we were unable to determine if participants drank tea with their meals or at other times, and tea drinking included both black and green tea.^60^ Another limitation is the modest representation of iron overload in the UK Biobank, with most participants having normal or borderline elevated levels, potentially limiting the study’s ability to investigate the full spectrum of iron loading.

## Conclusions

Male and female C282Y homozygotes without a hemochromatosis diagnosis exhibit excess liver iron, whereas other *HFE* genotypes show a low penetrance of iron loading. Liver iron levels are associated with genetic factors, alcohol consumption, smoking, body composition, PPI medication, and dietary habits. C282Y homozygotes diagnosed with hemochromatosis have significantly lower liver iron levels, likely due to treatment effects and lifestyle modifications. Our findings suggest that individuals at risk of hemochromatosis may benefit from tailored advice on lifestyle, diet and treatment to manage iron accumulation.

## Data Availability

All data produced are available online at the UK Biobank website

https://www.ukbiobank.ac.uk/

## Data sharing

Data are available on application to the UK Biobank (www.ukbiobank.ac.uk/register-apply).

## Abbreviations

+/+: Homozygous for a particular gene or mutation
+/-: Heterozygous for a particular gene or mutation
ALT: Alanine aminotransferase
AST: Aspartate aminotransferase
BMI: Body mass index
CSEs: Certificate of Secondary Education
GCSEs: General Certificate of Secondary Education
GWAS: Genome-wide association study
HC: Hip circumference
HbA1c: Glycated hemoglobin
*HFE*: Gene encoding *HFE* protein (Homeostatic iron regulator)
HNC: Higher National Certificate
HND: Higher National Diploma
LSIR: Liver-to-spleen iron ratio
MET: Metabolic equivalent task
mg/g: Milligrams per gram
mmol/L: Millimoles per liter
MRI: Magnetic resonance imaging
MRLIC: Magnetic resonance liver iron concentration
NVQ: National Vocational Qualification
PDFF: Proton density fat fraction
PGS: Polygenic score
PPI: Proton pump inhibitor
SF: Serum ferritin
TSAT: Transferrin saturation
UKB: UK Biobank
WC: Waist circumference
WHR: Waist-to-hip ratio
WHtR: Waist-to-height ratio

## Acknowledgements

This research was conducted using the UK Biobank resource, under application 14631. We thank the UK Biobank participants and coordinators for the dataset. We express our sincere gratitude to Emeritus Professor David Melzer for his invaluable guidance, support, and for providing the initial research idea that made this work possible. This work used data provided by patients and collected by the NHS as part of their care and support. Copyright © (2023), NHS England. Re-used with the permission of the NHS England and UK Biobank. All rights reserved. This study was supported by the National Institute for Health and Care Research Exeter Biomedical Research Centre. The views expressed are those of the authors and not necessarily those of the NIHR or the Department of Health and Social Care.

## Funding

JA has an NIHR Advanced Fellowship (NIHR301844). The University of Exeter supports ML, LP, JCD and DSW.

## Conflict of interest

None declared.

## Notes

### Competing Interest Statement

The authors have declared no competing interest.

### Funding Statement

The University of Exeter and NIHR

