## Supplementary Material for "Liver iron levels are associated with *HFE*-hemochromatosis genotype, diet, adiposity, and disease in the UK Biobank"

**eTable 1. Characteristics of UK Biobank participants by *HFE* genotype status, without a diagnosis of hemochromatosis**

| | EUR participants | EUR without C282Y and H63D variants | EUR *HFE* variants excl C282Y homozygotes | EUR C282Y homozygotes | Non-EUR participants |
|---|---|---|---|---|---|
| **n** | 37,229 | 22,388 | 14,750 | 91 | 2,859 |
| **MRI data** | | | | | |
| Liver iron (mg/g), mean (SD) | 1.27 (0.2) | 1.25 (0.2) | 1.31 (0.3) | 2.37 (1.2) | 1.25 (0.2) |
| Liver PDFF (%), mean (SD) | 4.89 (4.9) | 4.78 (4.8) | 4.82 (4.9) | 4.19 (4.0) | 4.92 (4.8) |
| Pancreas iron (mg/g), mean (SD) | 0.78 (0.1) | 0.78 (0.1) | 0.78 (0.1) | 0.78 (0.1) | 0.77 (0.1) |
| **Iron PGS** | | | | | |
| PGS TSAT (excl *HFE*), mean (SD) | 0.40 (0.1) | 0.32 (0.1) | 0.32 (0.1) | 0.31 (0.1) | 0.40 (0.1) |
| PGS SF (excl *HFE*), mean (SD) | 1.10 (0.1) | 1.10 (0.1) | 1.10 (0.1) | 1.09 (0.1) | 1.08 (0.1) |
| **Body composition** | | | | | |
| WHR ≥0.96 (males), n (%) | 5,668 (32.97) | 3,356 (32.38) | 2,301 (33.98) | 11 (47.83) | 388 (30.12) |
| WHR ≥0.85 (females), n (%) | 5,950 (31.60) | 3,519 (31.16) | 2,413 (32.29) | 18 (27.69) | 487 (33.15) |
| BMI groups (kg/m²), n (%) | | | | | |
| Underweight | 270 (0.75) | 158 (0.73) | 109 (0.77) | <5 | 19 (0.69) |
| Normal | 14,488 (40.33) | 8,762 (40.51) | 5,682 (40.00) | 44 (50.00) | 1,091 (39.66) |
| Overweight | 14,862 (41.37) | 8,940 (41.34) | 5,889 (41.46) | 32 (36.36) | 1,118 (40.64) |
| Obese | 6,302 (17.54) | 3,768 (17.42) | 2,525 (17.78) | 9 (10.23) | 523 (19.01) |
| WHtR >0.50, n (%) | 22,364 (64.84) | 13,324 (64.28) | 8,992 (65.76) | 47 (55.29) | 1,726 (65.26) |
| **Biomarkers** | | | | | |
| HbA1c (mmol/mol), mean (SD) | 34.99 (5.1) | 35.16 (5.1) | 34.03 (5.2) | 33.03 (3.8) | 36.14 (6.3) |
| Total cholesterol (mmol/L), mean (SD) | 5.74 (1.1) | 5.75 (1.1) | 5.71 (1.1) | 5.52 (1.0) | 5.65 (1.1) |
| Triglycerides (mmol/L), mean (SD) | 1.64 (1.0) | 1.64 (1.0) | 1.65 (1.0) | 1.51 (0.8) | 1.63 (1.0) |
| **Disease status** | | | | | |
| Viral hepatitis, n (%) | 89 (0.24) | 60 (0.27) | 29 (0.20) | 0 (0.00) | 14 (0.49) |
| Hypertension, n (%) | 7,720 (20.74) | 4,485 (20.03) | 3,234 (21.90) | 31 (24.80) | 635 (22.24) |
| Type 1 Diabetes, n (%) | 113 (0.30) | 65 (0.29) | 50 (0.34) | 0 (0.00) | 3 (0.11) |

| | | | | | |
|---|---|---|---|---|---|
| Type 2 Diabetes, n (%) | 981 (2.63) | 561 (2.51) | 418 (2.83) | 7 (5.60) | 123 (4.31) |
| **Vitamins/Minerals** | | | | | |
| Iron supplement use, n (%) | 1,174 (3.17) | 743 (3.34) | 428 (2.92) | <5 | 153 (5.41) |
| Vitamin C use, n (%) | 2,690 (7.27) | 1,642 (7.38) | 1,038 (7.08) | 10 (11.24) | 242 (8.55) |
| **Medication** | | | | | |
| Proton pump inhibitors, n (%) | 4,467 (12.00) | 2,670 (11.93) | 1,784 (12.09) | 13 (14.29) | 300 (10.51) |
| **Diet** | | | | | |
| Alcohol intake (units/week), n (%) | | | | | |
| 0 units/week | 3,723 (10.15) | 2,242 (10.17) | 1,471 (10.11) | 10 (11.49) | 565 (20.24) |
| 1–14 units/week | 20,521 (55.94) | 12,276 (55.63) | 8,199 (56.37) | 55 (63.22) | 1,614 (57.83) |
| 15–29 units/week | 8,603 (23.45) | 5,243 (23.78) | 3,343 (22.99) | 16 (18.39) | 423 (15.16) |
| ≥30 units/week | 3,837 (10.46) | 2,300 (10.43) | 1,531 (10.53) | 6 (6.90) | 189 (6.77) |
| Red/processed meat consumption (times/week), n (%) | | | | | |
| 0 times/week | 2,824 (7.59) | 1,708 (7.63) | 1,099 (7.45) | 17 (18.89) | 321 (11.24) |
| 0.1–2.9 times/week | 16,090 (43.22) | 9,681 (43.24) | 6,366 (43.16) | 42 (46.67) | 1,378 (48.27) |
| ≥3.0 times/week | 18,316 (49.20) | 10,999 (49.13) | 7,285 (49.39) | 32 (35.16) | 1,156 (40.49) |
| Tea intake (cups/day), n (%) | | | | | |
| 0 cups/day | 5,340 (14.45) | 3,185 (14.33) | 2,144 (14.64) | 11 (12.36) | 391 (13.83) |
| <1 cup/day | 1,052 (2.85) | 637 (2.87) | 410 (2.80) | 5 (5.62) | 125 (4.42) |
| 1–3 cups/day | 14,532 (39.32) | 8,752 (39.38) | 5,739 (39.18) | 40 (44.94) | 1,316 (46.55) |
| ≥4 cups/day | 16,037 (43.39) | 9,648 (43.42) | 6,356 (43.39) | 33 (37.08) | 995 (35.20) |
| **Lifestyle** | | | | | |
| Current smoker, n (%) | 1,219 (3.30) | 705 (3.17) | 512 (3.49) | <5 | 69 (2.48) |

Numbers are shown as mean (SD) for continuous variables and n (%) for categorical variables.

**Abbreviations:** BMI, body mass index; excl, excluding; HbA1c, hemoglobin A1c; MRLIC, magnetic resonance liver iron concentration; MRI, magnetic resonance imaging; mmol/L, millimoles per liter; PDFF, proton density fat fraction; PGR, polygenic score; SD, standard deviation; SF, serum ferritin; TSAT, transferrin saturation; WHR, waist-to-hip ratio; WHtR, waist-to-height ratio. PGS for transferrin saturation and serum ferritin were derived from a published GWAS.[2]

**eTable 2. Associations between environmental and genetic variables and magnetic resonance liver iron concentration in UK Biobank by *HFE* genotype status, without a diagnosis of hemochromatosis**

| | EUR participants | | EUR without C282Y and H63D variants | | EUR *HFE* variants excl C282Y homozygotes | | EUR C282Y homozygotes | | Non-EUR participants | |
|---|---|---|---|---|---|---|---|---|---|---|
| | β (95% CI) | p | β (95% CI) | p | β (95% CI) | p | β (95% CI) | p | β (95% CI) | p |
| **Iron PGS** | | | | | | | | | | |
| PGS TSAT (excl *HFE*) | 0.22 (0.19-0.26) | 3.80E-46 | 0.18 (0.15-0.21) | 9.90E-29 | 0.33 (0.27-0.38) | 1.10E-30 | -2.88 (-6.45-0.69) | 0.11 | 0.11 (-0.006-0.22) | 7.00E-02 |
| PGS SF (excl *HFE*) | 0.18 (0.15-0.21) | 2.10E-27 | 0.17 (0.14-0.20) | 1.60E-25 | 0.20 (0.14-0.25) | 3.80E-11 | 1.03 (-2.68-4.73) | 0.58 | 0.13 (0.02-0.25) | 3.00E-02 |
| **Body composition** | | | | | | | | | | |
| WHR males ≥0.96 | -0.004 (-0.01 - -0.004) | 0.33 | -0.008 (-0.02-0.0007) | 0.07 | -0.007 (-0.02-0.009) | 0.38 | 1.08 (-0.79-2.96) | 0.23 | -0.02 (-0.05-0.02) | 0.33 |
| WHR females ≥0.85 | 0.009 (0.002-0.02) | 9.60E-03 | 0.01 (0.004-0.02) | 0.003 | 0.005 (-0.006-0.02) | 0.37 | 0.03 (-0.67-0.73) | 0.93 | 0.02 (-0.008-0.04) | 0.18 |
| BMI | | | | | | | | | | |
| Underweight | -0.06 (-0.09 - -0.03) | 1.10E-04 | -0.05 (-0.08 - -0.02) | 0.001 | -0.08 (-0.13 - -0.03) | 0.003 | 0.44 (-1.15-2.02) | 0.59 | -0.03 (-0.14-0.08) | 0.57 |
| Normal | Reference | | Reference | | Reference | | Reference | | Reference | |
| Overweight | 0.02 (0.02-0.03) | 4.30E-15 | 0.01 (0.006-0.02) | 6.40E-05 | 0.02 (0.01-0.03) | 4.70E-05 | -0.004 (-0.66-0.66) | 0.99 | 0.02 (-0.002-0.04) | 0.07 |
| Obese | 0.01 (0.003-0.02) | 7.00E-03 | 0.009 (0.002-0.02) | 0.01 | 0.003 (-0.01-0.02) | 0.68 | -0.21 (-1.18-0.76) | 0.67 | 0.01 (-0.01-0.04) | 0.3 |
| WHtR >0.50 | 0.01 (0.006-0.02) | 6.40E-05 | 0.009 (0.003-0.01) | 2.80E-03 | 0.01 (0.003-0.02) | 8.90E-03 | 0.24 (-0.38-0.86) | 0.44 | 0.03 (0.008-0.05) | 7.00E-03 |
| **Biomarkers** | | | | | | | | | | |
| HbA1c | -0.004 (-0.004 - -0.003) | 4.10E-48 | -0.002 (-0.003 - -0.002) | 3.00E-17 | -0.004 (-0.005 - -0.004) | 1.10E-21 | -0.08 (-0.16-0.001) | 0.05 | -0.002 (-0.003 - - 0.003) | 0.02 |
| Cholesterol | 0.009 (0.007-0.01) | 1.30E-13 | 0.009 (0.007-0.01) | 1.10E-13 | 0.01 (0.007-0.02) | 2.30E-07 | 0.30 (-0.02-0.62) | 0.07 | 0.004 (-0.004-0.01) | 0.35 |
| Triglycerides | 0.006 (0.003-0.009) | 1.20E-05 | 0.003 (-0.0001-0.006) | 0.06 | 0.01 (0.006-0.02) | 8.00E-06 | 0.008 (-0.41-0.43) | 0.97 | 0.03 (-0.007-0.01) | 0.51 |
| **Diseases** | | | | | | | | | | |
| Viral hepatitis | -0.007 (-0.06-0.04) | 0.78 | 0.03 (-0.02-0.07) | 0.32 | -0.05 (-0.16-0.05) | 0.3 | N/A | N/A | 0.01 (-0.012-0.15) | 0.83 |
| Hypertension | -0.005 (-0.01-0.002) | 0.14 | 0.001 (-0.005-0.008) | 0.75 | -0.01 (-0.02-0.001) | 0.08 | -0.60 (-1.37-0.17) | 0.12 | -0.03 (-0.05 - -0.003) | 0.03 |
| Type 1 Diabetes | -0.08 (-0.12 - -0.03) | 9.90E-04 | -0.07 (-0.12 - -0.02) | 0.003 | -0.09 (-0.17 - -0.008) | 0.31 | N/A | N/A | 0.56 (0.17-0.96) | 0.005 |
| Type 2 Diabetes | -0.07 (-0.08 - -0.05) | 8.20E-17 | -0.05 (-0.07 - -0.04) | 2.10E-10 | -0.08 (-0.11 - -0.05) | 3.90E-09 | -1.24 (-2.62-0.13) | 0.08 | -0.05 (-0.09 - -0.01) | 0.02 |
| **Vitamins/Minerals** | | | | | | | | | | |
| Iron supplement | -0.03 (-0.05 - -0.02) | 7.60E-06 | -0.02 (-0.04 - -0.008) | 0.002 | -0.04 (-0.07 - -0.02) | 0.01 | -0.67 (-2.16-0.81) | 0.37 | -0.03 (-0.07-0.006) | 0.1 |
| Vitamin C | -0.009 (-0.02-0.0004) | 0.06 | -0.004 (-0.01-0.006) | 0.45 | -0.02 (-0.03-0.003) | 0.09 | -0.62 (-1.46-0.22) | 0.15 | -0.02 (-0.06-0.008) | 0.14 |
| **Medication** | | | | | | | | | | |
| Proton pump inhibitors | -0.03 (-0.04 - -0.03) | 3.50E-17 | -0.02 (-0.03 - -0.01) | 4.10E-08 | -0.05 (-0.06 - -0.03) | 7.70E-12 | -0.07 (-0.90-0.76) | 0.87 | -0.02 (-0.05-0.01) | 0.28 |
| **Diet** | | | | | | | | | | |
| Alcohol intake | | | | | | | | | | |

| | | | | | | | | | | |
|---|---|---|---|---|---|---|---|---|---|---|
| 0 units per week | -0.02 (-0.03 - -0.01) | 3.30E-07 | -0.02 (-0.02 - -0.007) | 3.10E-04 | -0.03 (-0.05 - -0.02) | 2.20E-05 | 0.33 (-0.55-1.21) | 0.46 | -0.01 (-0.04-0.008) | 0.22 |
| 1-14 units per week | Reference | | Reference | | Reference | | Reference | | Reference | |
| 15-29 units per week | 0.05 (0.05-0.06) | 7.80E-65 | 0.05 (0.04-0.06) | 1.80E-53 | 0.06 (0.05-0.07) | 2.50E-26 | 0.55 (-0.21-1.31) | 0.15 | 0.04 (0.01-0.07) | 0.007 |
| ≥30 units per week | 0.11 (0.10-0.12) | 6.00E-128 | 0.09 (0.08-0.10) | 7.20E-86 | 0.13 (0.12-0.15) | 1.00E-60 | 1.07 (-0.16-2.30) | 0.09 | 0.09 (0.05-0.13) | 1.70E-05 |
| Red/processed meat consumption | | | | | | | | | | |
| 0 times/week | Reference | | Reference | | Reference | | Reference | | Reference | |
| 0.1-2.9 times/week | 0.05 (0.04-0.06) | 1.90E-20 | 0.04 (0.03-0.05) | 1.30E-12 | 0.07 (0.05-0.08) | 2.20E-13 | 0.42 (-0.33-1.17) | 0.27 | 0.04 (0.01-0.07) | 4.40E-03 |
| ≥3.0 times/week | 0.08 (0.07-0.09) | 3.70E-54 | 0.06 (0.05-0.07) | 2.20E-35 | 0.11 (0.09-0.12) | 2.60E-31 | 0.56 (-0.29-1.40) | 0.19 | 0.07 (0.04-0.10) | 1.80E-06 |
| Tea intake per day | | | | | | | | | | |
| 0 cups per day | Reference | | Reference | | Reference | | Reference | | Reference | |
| <1 cup/day | 0.007 (-0.009-0.02) | 0.38 | 0.008 (-0.009-0.02) | 0.37 | 0.003 (-0.03-0.03) | 0.83 | -0.68 (-2.04-0.68) | 0.32 | -0.009 (-0.06-0.04) | 0.71 |
| 1-3 cups/day | -0.006 (-0.01-0.002) | 0.15 | -0.006 (-0.01-0.002) | 0.12 | -0.001 (-00.02-0.01) | 0.86 | -0.91 (-1.76 - -0.05) | 0.04 | 0.004 (-0.02-0.03) | 0.78 |
| ≥4 cups/day | -0.01 (-0.02 - -0.003) | 0.005 | -0.007 (-0.01-0.0007) | 0.07 | -0.01 (-0.03-0.002) | 0.1 | -0.99 (-1.90 - -0.09) | 0.03 | -0.007 (-0.04-0.02) | 0.65 |
| **Lifestyle** | | | | | | | | | | |
| Current smoker | 0.03 (0.02-0.04) | 5.10E-05 | 0.008 (-0.006-0.02) | 0.26 | 0.06 (0.03-0.08) | 6.80E-06 | 0.21 (-1.65-2.10) | 0.83 | 0.04 (-0.02-0.10) | 0.17 |

Linear regression beta-coefficients and 95% confidence intervals. Models were adjusted for age, sex, and 10 genetic principal components (PCs) to reduce bias from population stratification.

**Abbreviations:** BMI, body mass index; CI, confidence intervals; excl, excluding; HbA1c, hemoglobin A1c; MRLIC, magnetic resonance liver iron concentration; MRI, magnetic resonance imaging; mmol/L, millimoles per liter; PDFF, proton density fat fraction; PGR, polygenic score; p-value, probability value; SD, standard deviation; SF, serum ferritin; TSAT, transferrin saturation; WHR, waist-to-hip ratio; WHtR, waist-to-height ratio; β, beta-coefficients. PGS for transferrin saturation and serum ferritin were derived from a published GWAS.[2]

**eTable 3. Adjusted models: Associations between environmental and genetic variables and magnetic resonance liver iron concentration in UK Biobank by *HFE* genotype status, without a diagnosis of hemochromatosis**

| | EUR participants | | EUR without C282Y and H63D variants | | EUR *HFE* variants excl C282Y homozygotes | | EUR C282Y homozygotes | | Non-EUR participants | |
|---|---|---|---|---|---|---|---|---|---|---|
| | β (95% CI) | p | β (95% CI) | p | β (95% CI) | p | β (95% CI) | p | β (95% CI) | p |
| **Iron PGS** | | | | | | | | | | |
| PGS TSAT (excl *HFE*) | 0.23 (0.20-0.26) | 7.60E-47 | 0.18 (0.15-0.21) | 2.80E-30 | 0.18 (-1.99-2.35) | 0.86 | -3.96 (-8.48-0.55) | 0.08 | 0.11 (-0.003-0.23) | 0.06 |
| PGS SF (excl *HFE*) | 0.18 (0.15-0.21) | 1.90E-27 | 0.16 (0.13-0.20) | 1.90E-23 | 0.20 (0.14-0.26) | 1.40E-11 | 2.87 (-1.30-7.04) | 0.17 | 0.16 (0.04-0.29) | 0.009 |
| **Body composition** | | | | | | | | | | |
| WHR males ≥0.96 | -0.002 (-0.01-0.007) | 0.71 | -0.005 (-0.01-0.004) | 0.29 | -0.006 (-0.02-0.01) | 0.49 | N/A | N/A | -0.006 (-0.04-0.03) | 0.73 |
| WHR females ≥0.85 | 0.01 (0.006-0.02) | 0.0005 | 0.01 (0.006-0.02) | 0.0002 | 0.009 (-0.003-0.02) | 0.13 | 0.007 (-0.90-0.91) | 0.99 | 0.01 (-0.01-0.04) | 0.34 |
| BMI | | | | | | | | | | |
| Underweight | -0.03 (-0.06 - -0.003) | 0.03 | -0.04 (-0.07 - -0.007) | 0.02 | -0.06 (-0.11-0.01) | 0.02 | 0.87 (-1.16-2.90) | 0.39 | -0.03 (-0.14-0.08) | 0.61 |
| Normal | Reference | | Reference | | Reference | | Reference | | Reference | |
| Overweight | 0.008 (0.002-0.01) | 0.01 | 0.005 (-0.001-0.01) | 0.12 | 0.01 (0.004-0.03) | 0.006 | 0.02 (-0.95-0.99) | 0.97 | 0.01 (-0.009-0.04) | 0.26 |
| Obese | -0.002 (-0.01-0.005) | 0.49 | 0.003 (-0.005-0.01) | 0.5 | -0.003 (-0.02-0.01) | 0.69 | -0.60 (-2.05-0.84) | 0.41 | 0.009 (-0.02-0.04) | 0.54 |
| WHtR >0.50 | 0.02 (0.009-0.02) | 1.70E-05 | 0.01 (0.005-0.02) | 7.50E-04 | 0.02 (0.01-0.04) | 6.10E-04 | 0.38 (-0.82-1.57) | 0.53 | 0.04 (0.01-0.06) | 4.40E-03 |
| **Biomarkers** | | | | | | | | | | |
| HbA1c | -0.003 (-0.004 - -0.002) | 1.80E-30 | -0.002 (-0.003 - -0.001) | 8.30E-11 | -0.004 (-0.005 - -0.003) | 4.30E-13 | -0.08 (-0.20-0.04) | 0.17 | -0.002 (-0.003-0.0002) | 0.08 |
| Total cholesterol | 0.005 (0.003-0.007) | 7.10E-05 | 0.006 (0.003-0.008) | 5.40E-06 | 0.006 (0.002-0.01) | 0.007 | -0.008 (-0.43-0.41) | 0.97 | -0.0004 (-0.009-0.009) | 0.93 |
| Triglycerides | 0.009 (0.006-0.01) | 4.40E-09 | 0.004 (0.0007-0.007) | 0.02 | 0.02 (0.01-0.02) | 7.90E-10 | -0.13 (-0.61-0.36) | 0.61 | 0.003 (-0.008-0.01) | 0.62 |
| **Disease status** | | | | | | | | | | |
| Viral hepatitis | 0.003 (-0.05-0.05) | 0.91 | 0.04 (-0.01-0.09) | 0.13 | -0.04 (-0.14-0.06) | 0.42 | N/A | N/A | 0.01 (-0.13-0.15) | 0.87 |
| Hypertension | -0.002 (-0.008-0.005) | 0.6 | 0.002 (-0.005-0.009) | 0.56 | -0.005 (-0.02-0.006) | 0.35 | -0.35 (-1.37-0.67) | 0.49 | -0.03 (-0.06 - -0.007) | 0.01 |
| Type 1 Diabetes | -0.06 (-0.11 - -0.01) | 0.01 | -0.06 (-0.10 - -0.007) | 0.02 | -0.07 (-0.15-0.01) | 0.09 | N/A | N/A | 0.62 (0.19-1.04) | 0.004 |
| Type 2 Diabetes | -0.06 (-0.08-0.05) | 6.50E-15 | -0.05 (-0.07 - -0.04) | 9.80E-10 | -0.07 (-0.10-0.05) | 1.60E-07 | 1.24 (-2.94-0.46) | 0.15 | -0.05 (-0.10 - -0.006) | 0.03 |
| **Vitamins/Minerals** | | | | | | | | | | |
| Iron supplement | -0.02 (-0.03 - -0.004) | 0.01 | -0.007 (-0.002-0.007) | 0.31 | -0.03 (-0.06 - -0.001) | 0.04 | 0.51 (-1.20-2.22) | 0.55 | -0.01 (-0.05-0.03) | 0.53 |
| Vitamin C | -0.004 (-0.01-0.006) | 0.48 | -0.0001 (-0.01-0.01) | 0.98 | -0.007 (-0.02-0.01) | 0.45 | -0.86 (-1.92-0.20) | 0.11 | -0.02 (-0.05-0.01) | 0.23 |
| **Medication** | | | | | | | | | | |
| Proton pump inhibitors | -0.03 (-0.04 - -0.03) | 1.40E-18 | -0.02 (-0.03 - -0.02) | 6.40E-10 | -0.05 (-0.06 - -0.04) | 6.50E-13 | 0.01 (-0.93-0.96) | 0.98 | -0.01 (-0.05-0.02) | 0.37 |

| | | | | | | | | | | |
|---|---|---|---|---|---|---|---|---|---|---|
| **Diet** | | | | | | | | | | |
| Alcohol intake | | | | | | | | | | |
| 0 units/week | -0.02 (-0.03 - -0.009) | 9.40E-05 | -0.01 (-0.02 - -0.003) | 9.80E-03 | -0.03 (-0.04 - -0.01) | 0.001 | 0.21 (-0.74-1.16) | 0.66 | -0.01 (-0.03-0.01) | 0.38 |
| 1-14 units/week | Reference | | Reference | | Reference | | Reference | | Reference | |
| 15-29 units/week | 0.05 (0.04-0.06) | 4.40E-54 | 0.05 (0.04-0.05) | 2.70E-46 | 0.06 (0.05-0.07) | 4.40E-23 | 0.29 (-0.52-1.10) | 0.48 | 0.04 (0.005-0.07) | 0.02 |
| ≥30 units/week | 0.10 (0.09-0.11) | 5.00E-109 | 0.08 (0.07-0.09) | 1.10E-71 | 0.13 (0.11-0.14) | 9.50E-55 | 1.13 (-0.43-2.68) | 0.15 | 0.09 (0.04-0.13) | 7.90E-05 |
| Red/processed meat consumption | | | | | | | | | | |
| 0 times/week | Reference | | Reference | | Reference | | Reference | | Reference | |
| 0.1-2.9 times/week | 0.05 (0.04-0.06) | 8.30E-21 | 0.04 (0.03-0.05) | 7.60E-12 | 0.07 (0.05-0.09) | 7.30E-13 | 1.09 (0.05-2.12) | 0.41 | 0.05 (0.02-0.08) | 0.003 |
| ≥3.0 times/week | 0.07 (0.06-0.09) | 2.40E-45 | 0.06 (0.05-0.07) | 4.50E-28 | 0.10 (0.08-0.12) | 7.50E-26 | 1.13 (0.06-2.21) | 0.04 | 0.07 (0.04-0.11) | 1.10E-05 |
| Tea intake per day | | | | | | | | | | |
| 0 cups/day | Reference | | Reference | | Reference | | Reference | | Reference | |
| <1 cup/day | 0.005 (-0.01-0.02) | 0.54 | 0.006 (-0.01-0.02) | 0.49 | 0.001 (-0.03-0.03) | 0.95 | -1.13 (-2.68-0.41) | 0.15 | -0.007 (-0.06-0.04) | 0.78 |
| 1-3 cups/day | -0.008 (-0.02 - -0.0002) | 0.04 | -0.008 (-0.02-0.0003) | 0.06 | -0.008 (-0.02-0.006) | 0.29 | -1.03 (-2.02 - -0.05) | 0.04 | 0.008 (-0.02-0.04) | 0.57 |
| ≥4 cups/day | -0.01 (-0.02 - -0.005) | 1.20E-03 | -0.009 (-0.02 - -0.0008) | 0.03 | -0.02 (-0.03 - -0.003) | 0.02 | -0.81 (-1.83-0.21) | 0.12 | 0.003 (-0.03-0.03) | 0.85 |
| **Lifestyle** | | | | | | | | | | |
| Current smoker | 0.01 (-0.0006-0.03) | 0.06 | -0.004 (-0.02-0.01) | 0.59 | 0.04 (0.01-0.06) | 0.004 | 0.72 (-1.52-2.95) | 0.52 | 0.04 (-0.02-0.10) | 0.21 |

Linear regression beta-coefficients and 95% confidence intervals. Models were adjusted for age, sex, 10 principal components, assessment center, red/processed meat consumption, WHR, alcohol intake, PPI use, education, socioeconomic status, smoking, hepatitis, type 2 diabetes, tea drinking, iron and vitamin C supplement intake.

**Abbreviations:** BMI, body mass index; CI, confidence intervals; HbA1c, hemoglobin A1c; MRLIC, magnetic resonance liver iron concentration; MRI, magnetic resonance imaging; mmol/L, millimoles per liter; PDFF, proton density fat fraction; PGR, polygenic score; p, probability value; SD, standard deviation; SF, serum ferritin; TSAT, transferrin saturation; WHR, waist-to-hip ratio; WHtR, waist-to-height ratio; β, beta-coefficients. PGS for transferrin saturation and serum ferritin were derived from a published GWAS.[2]

**eTable 4. Associations between magnetic resonance liver iron concentration and risk of incident disease outcomes in UK Biobank by *HFE* genotype status, without a diagnosis of hemochromatosis**

| Outcome | EUR participants | EUR without *HFE* C282Y/H63D variants | C282Y homozygotes |
|---|---|---|---|
| **Incident N** | 37,229 | 22,388 | 91 |
| **Any liver disease**, n (%) | 442 (1.19%) | 230 (1.03%) | <5 |
| **Liver fibrosis/cirrhosis**, n (%) | 39 (0.10%) | 22 (0.10%) | <5 |
| **Hemochromatosis**, n (%) | 15 (0.04%) | 6 (0.03%) | 5 (4.00%) |
| **All-cause mortality**, n (%) | 670 (1.80%) | 390 (1.74%) | <5 |
| **Model 1:** | | | |
| Any liver disease (HR [95% CI], p) | 0.84 (0.56–1.26), p=0.40 | 1.12 (0.58–2.16), p=0.74 | N/A |
| Liver fibrosis/cirrhosis (HR [95% CI], p) | 0.10 (0.01–0.73), p=0.02 | 0.11 (0.007–1.87), p=0.13 | N/A |
| Hemochromatosis (HR [95% CI], p) | 5.40 (3.50–8.33), $p=2.6\times10^{-14}$ | 4.67 (0.38–58.11), p=0.23 | 0.79 (0.26–2.40), p=0.68 |
| **Model 2: Excluding any diagnosis of anaemia** | | | |
| N | 36,466 | 21,914 | 89 |
| Any liver disease (HR [95% CI], p) | 0.92 (0.61–1.39), p=0.70 | 1.34 (0.68–2.64), p=0.40 | N/A |
| Liver fibrosis/cirrhosis (HR [95% CI], p) | 0.11 (0.01–1.02), p=0.052 | 0.09 (0.003–2.80), p=0.17 | N/A |
| Hemochromatosis (HR [95% CI], p) | 5.78 (3.73–8.95), $p=3.8\times10^{-15}$ | 7.49 (0.81–68.95), p=0.08 | 0.68 (0.20–2.31), p=0.54 |
| **Model 3: Model 1 repeated + Adjusted (multivariable)** | | | |
| Any liver disease (HR [95% CI], p) | 1.05 (0.70–1.56), p=0.81 | 1.27 (0.64–2.54), p=0.50 | N/A |
| Liver fibrosis/cirrhosis (HR [95% CI], p) | 0.15 (0.02–1.13), p=0.07 | 0.13 (0.007–2.41), p=0.17 | N/A |
| Hemochromatosis (HR [95% CI], p) | 5.57 (3.52–8.81), $p=2.0\times10^{-13}$ | 3.23 (0.22–46.72), p=0.39 | N/A |

Cox proportional hazards models were used throughout. Models 1 and 2 – adjusted for age, sex, and 10 PCs. Model 3 – additionally adjusted for alcohol intake, smoking, BMI, and prevalent comorbidities: type 2 diabetes, any viral hepatitis, alcoholic liver disease, and NAFLD. **Abbreviations**: EUR, European; PCs, principal components; BMI, body mass index; NAFLD, non-alcoholic fatty liver disease; MRI, magnetic resonance imaging. MRLIC, MRI liver iron concentration.

**eFigure 1. Distribution of liver to spleen iron ratio by *HFE* genotype and hemochromatosis diagnosis status**

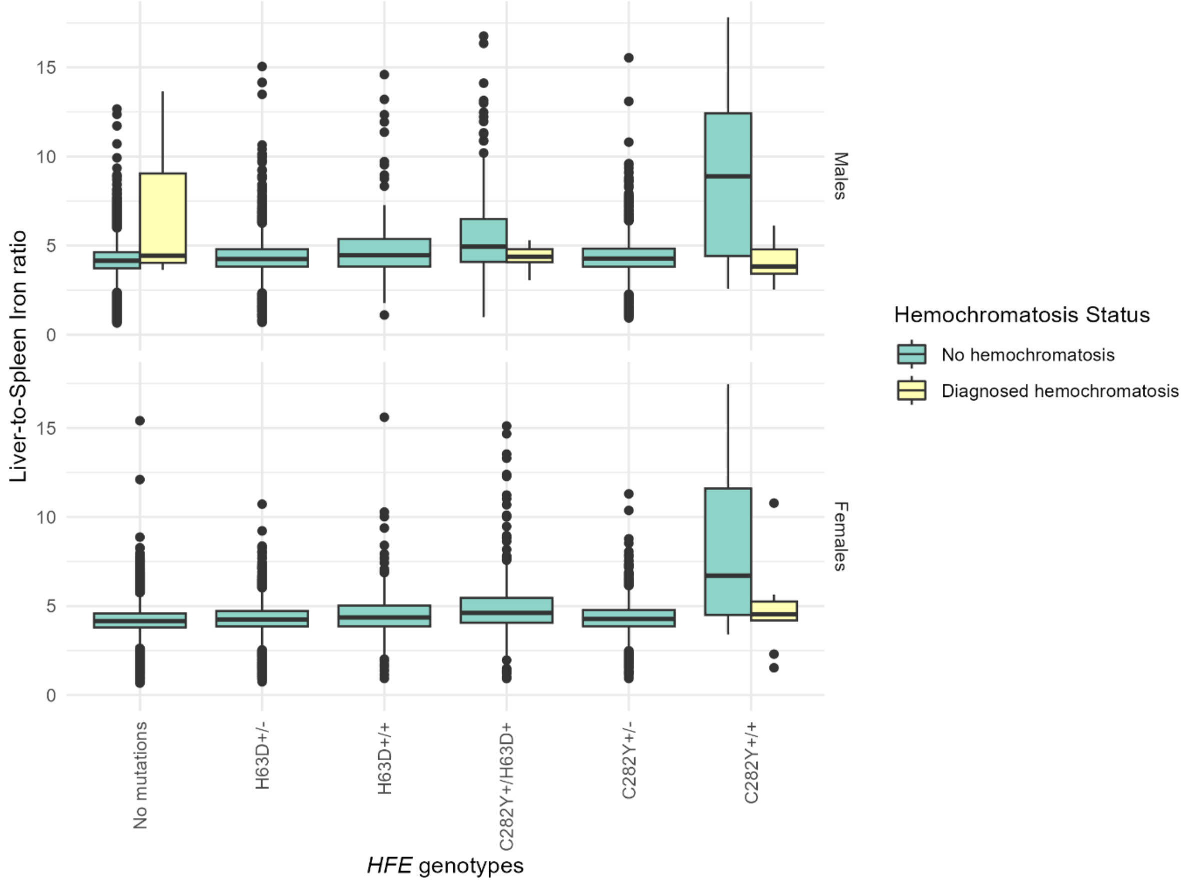

Distribution of MRI liver-spleen iron ratio (male n = 13,981; female n = 15,430) by *HFE* genotypes, stratified by hemochromatosis diagnosis, in the UK Biobank.

### eFigure 2. Distribution of liver PDFF by *HFE* genotype and hemochromatosis diagnosis status

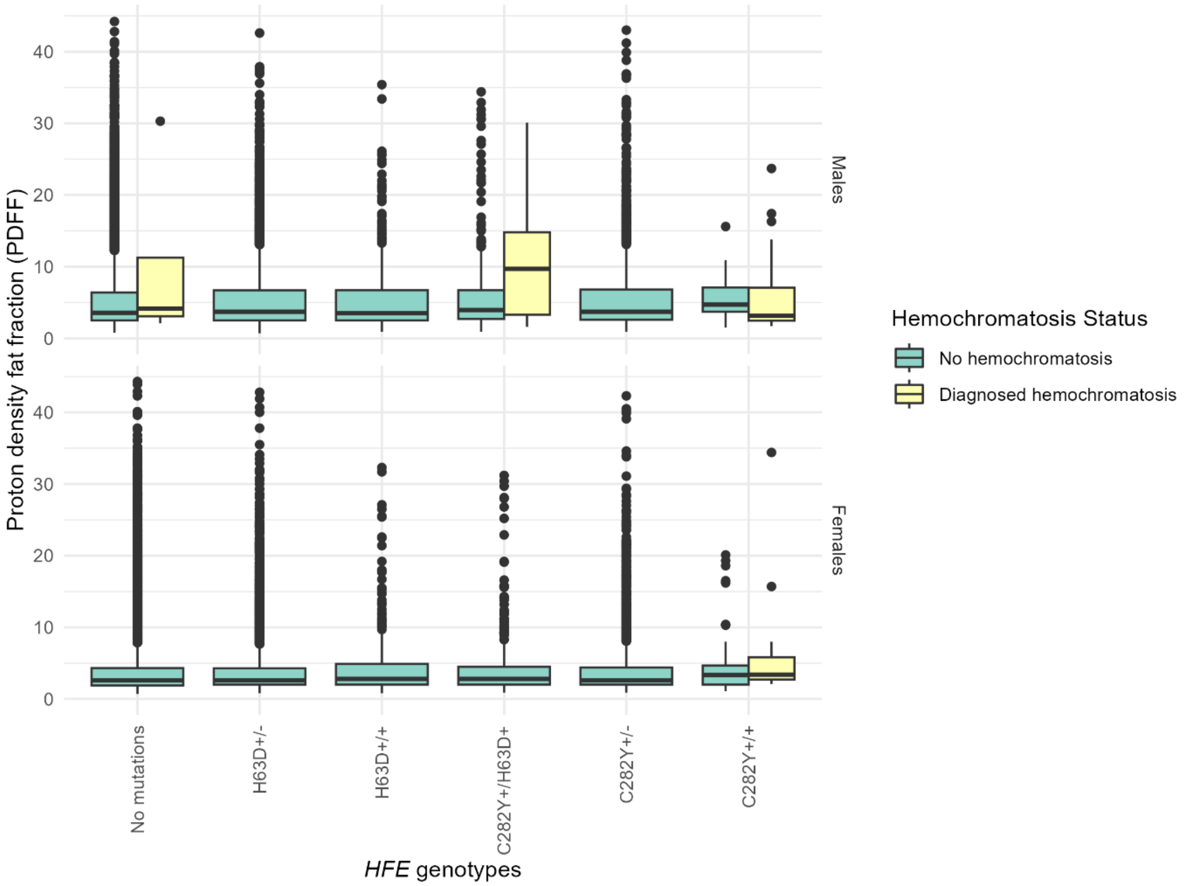

Distribution of MRI liver PDFF (%) (male n = 17,293; female n = 19,127) by *HFE* genotypes, stratified by hemochromatosis diagnosis, in the UK Biobank. Abbreviations: PDFF, proton density fat fraction.

### eFigure 3. Distribution of pancreas iron by *HFE* genotype and hemochromatosis diagnosis status

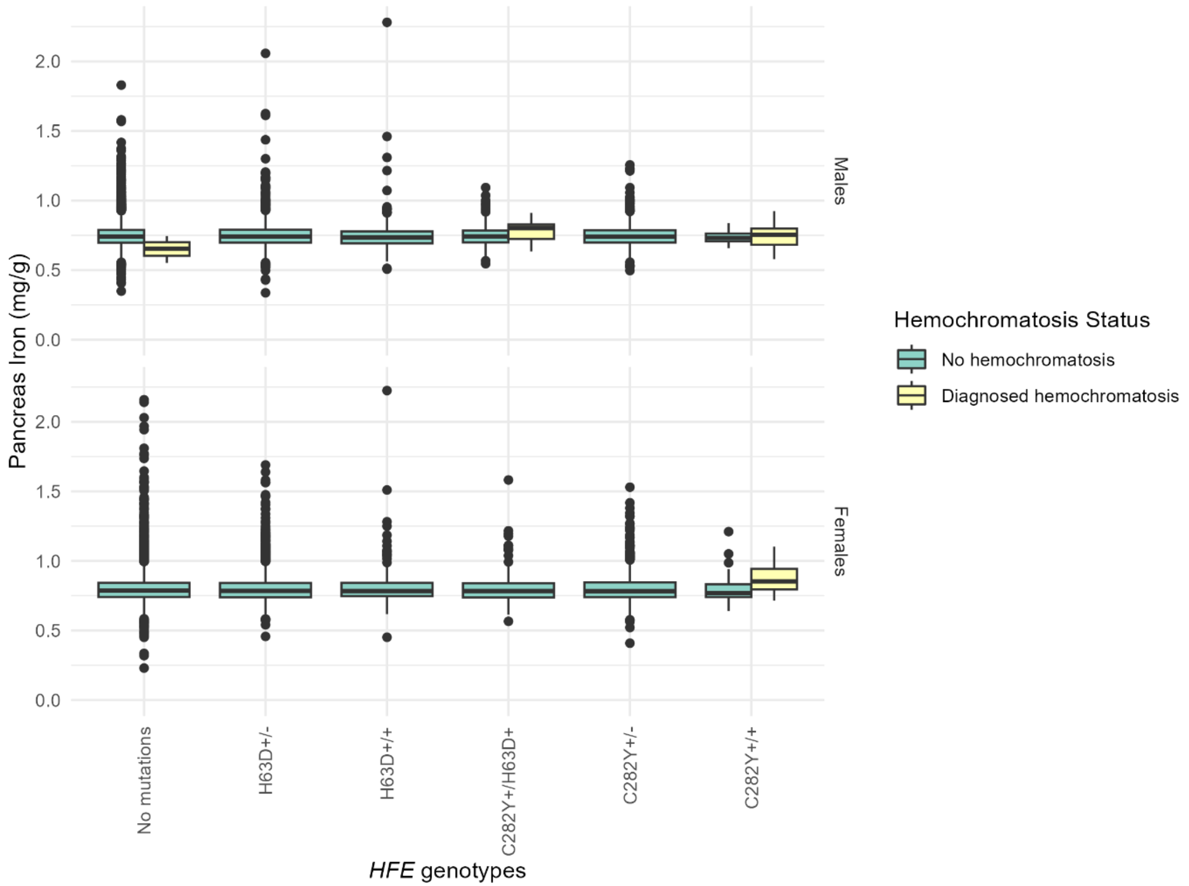

Distribution of MRI pancreas iron (male n = 12,424; female n = 13,483) by *HFE* genotypes, stratified by hemochromatosis diagnosis, in the UK Biobank. Abbreviations: mg/g, milligrams per gram.
